# Pneumococcal pneumonia in adults – Re-emergence of vaccine serotypes: a prospective multicentre cohort study in Germany, 2020-2023

**DOI:** 10.1101/2025.10.24.25338303

**Authors:** Christina Bahrs, Norman Rose, Grit Barten-Neiner, Carolin Fleischmann-Struzek, Mark P. G. van der Linden, Gernot Rohde, Jan Rupp, Martin Witzenrath, Jessica Rademacher, Mathias W. Pletz, the CAPNETZ Study Group

## Abstract

This prospective multicentre study investigated pneumococcal serotype distribution and vaccine coverage of the 13- and 20-valent conjugate vaccines, and the 23-valent polysaccharide vaccine among adults with community-acquired pneumonia in Germany, from 2020 to 2023, using serotype-specific urine antigen detection testing. The findings show that *Streptococcus pneumoniae* has reemerged as causative pathogen of pneumonia, particularly in adults aged ≥60 years. This resurgence, observed during the ongoing SARS-CoV-2 pandemic, has been primarily driven by an increase in cases due to serotype 3.

---

A previous CAPNETZ cohort study reported a marked shift in the aetiology of community-acquired pneumonia (CAP) among adults in Germany, with SARS-CoV-2 replacing common pathogens, such as *Streptococcus pneumoniae*, after April 2020 [1]. However, a prospective, population-based study in Thuringia (ThEpiCAP), Germany, found a re-emergence of *S. pneumoniae* as a causative pathogen in adults with CAP, beginning in the second half of 2021 [2]. Therefore, this raises the question, of whether the CAPNETZ cohort, that includes patients all over Germany, provides a more robust basis for evaluating the re-emergence of *S. pneumoniae* in Germany as a predominant etiological agent and the associated implications for pneumococcal vaccine coverage. Accordingly, this prospective multicentre study aimed to analyse pneumococcal serotype distribution and vaccine coverage of the 13-valent conjugate vaccine (PCV13), the 20-valent conjugate vaccine (PCV20), and the 23-valent polysaccharide vaccine (PPV23) among adults with CAP from 2020 to 2023. We analysed all patients enrolled in the CAPNETZ study (DRKS00005274, Ethics No. 104/01) between January 1, 2020, and December 31, 2023, at 26 centres in Germany. Participants gave written informed consent and provided urine samples for serotype-specific urine antigen detection (SSUAD) testing [3]. Eligible subjects were adults (≥18 years) with radiologically confirmed CAP and at least one clinical symptom. Exclusion criteria included hospital admission within the previous 28 days or new pulmonary tuberculosis diagnosed in the past two months. In addition to conventional pneumococcal urine antigen test (*BinaxNOW® S. pneumoniae*) and blood culture (BC) [4], we prospectively collected urine samples for SSUAD testing for detecting PCV13 serotypes [5] and 11 additional serotypes unique to PCV20 and PPV23 [6]. Samples were treated as previously described [7] and the analyses were performed at Pfizer’s Vaccines Research and Development Laboratory (Pearl River, NY, USA). Results were classified into “positive”, “indeterminate or missing” (both excluded from analysis) and “negative”. In accordance with STIKO’s 2016 recommendation for adult pneumococcal vaccination, individuals were classified as "at risk for pneumococcal disease" if they were aged ≥60, younger adults with a comorbidity, or severely immunosuppressed individuals [8]. We present the absolute and relative frequencies of PCV13, PCV20, and PPV23 pneumococcal vaccine serotypes stratified by year and patient group, along with cluster-robust confidence intervals (CIs). Annual trends from 2020–2023 were analysed using cluster-robust generalized linear models with heteroscedasticity-consistent standard errors. This approach accounts for data dependencies arising from the nested data structure (e.g., patients nested within hospitals). We used the saturated model g(Y_t_) = α + β_1_T_year > 2020_ + β_2_T_year > 2021_ + β_3_T_year= 2023_, with the logit-link function g(). Due to the time coding according to a change score model using the indicator variables T_j_, the regression coefficient β_j_ represents the log. odds ratios (OR) between two consecutive years. The trend was quantified by the average OR 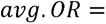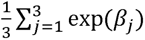. Standard errors and 95% CIs of the derived parameters avg.OR and avg.Δ were calculated with delta method [9]. All statistical analyses were performed using the R software including the survey and car packages [10].

Of the 2,028 patients with all-cause CAP, 1,504 (74.16%) had urine samples analysed with SSUAD tests. The majority were male (63.30%) and had at least one comorbidity (88.83%). Almost all received inpatient care (98.80%). The mean age was 65.5 years (range 18–99 years), with approximately one third younger than 60 years. Vaccination rates against influenza and pneumococcal diseases were 41.29% and 20.68%, respectively. The most frequent comorbidities were cardiovascular diseases (61.50%), chronic respiratory illnesses (36.17%), and malignancies (21.61%). Severe immunocompromise was identified in 8.64% of patients. Mortality was 6.25% after 28 days and increased to 11.17% by 180 days.

During the study period from 2020 to 2023, a total of 1,492 patients had valid SSUAD test results, of whom 131 (8.78%) patients tested positive. The SSUAD results were indeterminate in 7 (0.47%) patients, and data were missing for 5 (0.33%) patients. Among 1,482 patients tested with the BINAX NOW assay, 76 (5.13%) were positive, while 22 (1.46%) patients did not undergo the BINAX NOW test. *S. pneumoniae* was identified in BCs of 24 (2.42%) of the 990 patients who had at least one BC performed. Among patients with bacteraemic pneumococcal pneumonia, the following serotypes were detected by SSUAD: serotype 3 (n = 8, 33.33% of cases), serotype 8 (n = 3, 12.50%), serotype 4 (n = 1, 4.17%), serotype 10A (n = 1, 4.17%), serotype 19A (n = 1, 4.17%), serotype 20 (n = 1, 4.17%), and serotype 22F (n = 1, 4.17%). In 8 (33.33%) patients with bacteraemic pneumococcal CAP, no vaccine serotype could be identified by SSUAD.

Among the study cohort with all-cause CAP, serotype 3 was the most prevalent serotype, detected by SSUAD in 52 cases including the 8 patients with *S. pneumoniae* bacteraemia. This was followed by serotype 8 in 17 cases, serotype 22F in 8 cases, serotype 6A in 7 cases, and serotype 4 and 19A in 6 cases each. Serotypes 11A, 9N, and 20 were each found in 5 cases. Overall proportion of vaccine-type pneumococcal pneumonia among all-cause CAP for PCV13, PCV20, and PPV23 serotypes was 5.41% (95% CI 4.12-7.06%), 8.11% (95% CI 6.35-10.31%), and 8.31% (95% CI 6.27-10.93%), respectively. Significantly increasing annual trends were observed for pneumococcal pneumonia detected by BINAX NOW test (OR 1.77, 95% CI 1.37-2.17), for serotype 3 (OR 1.85, 95% 1.19-2.51), and for PPV23 serotypes (OR 1.35, 95% 1.03-1.67). Table 1 presents the proportions of pneumococcal serotypes among all-cause CAP from 2020 to 2023, along with the serotype distributions stratified according to the STIKO classifications. Serotype 3 was the most common serotype, followed by serotype 8, in both subgroups: those aged ≥60 years, and those aged 18-59 years with at least one comorbidity. Notably, among patients aged ≥60 years, an increasing annual trend was observed exclusively for serotype 3 (OR 1.84, 95% CI 1.01-2.67) and PPV23 serotypes (OR 1.47, 1.04-1.91). In the subgroup of 130 patients with severe immunosuppression, serotype 6A was the most prevalent, followed by serotypes 3 and 8.

**Table 1.**
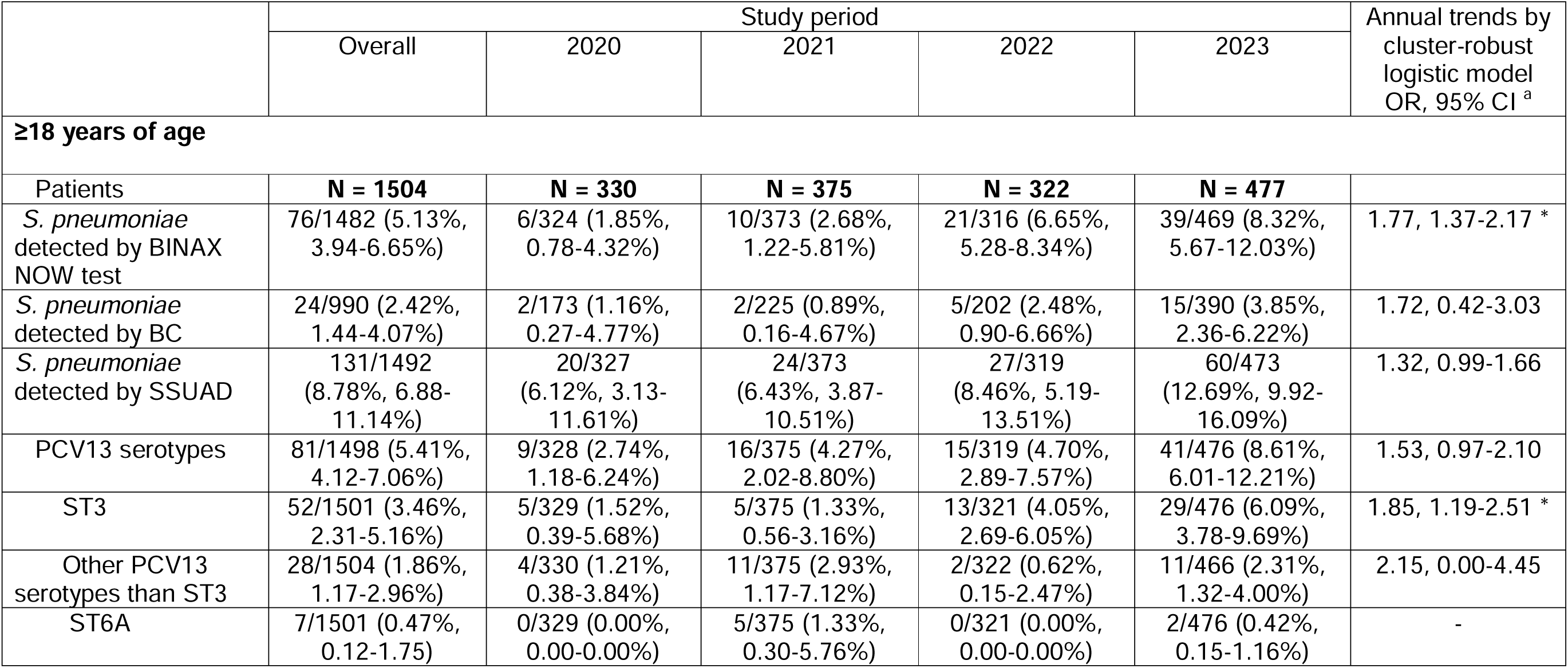
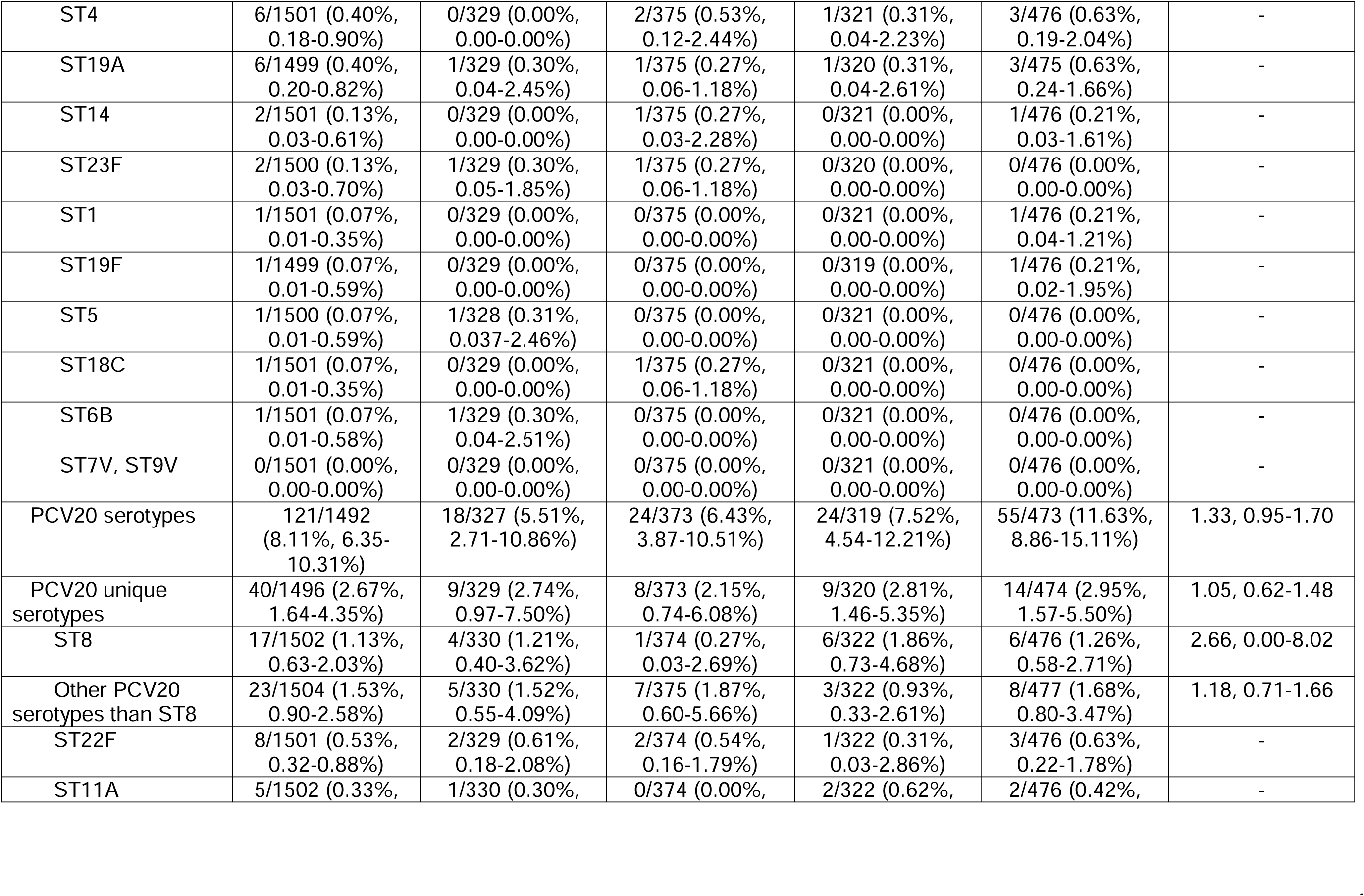
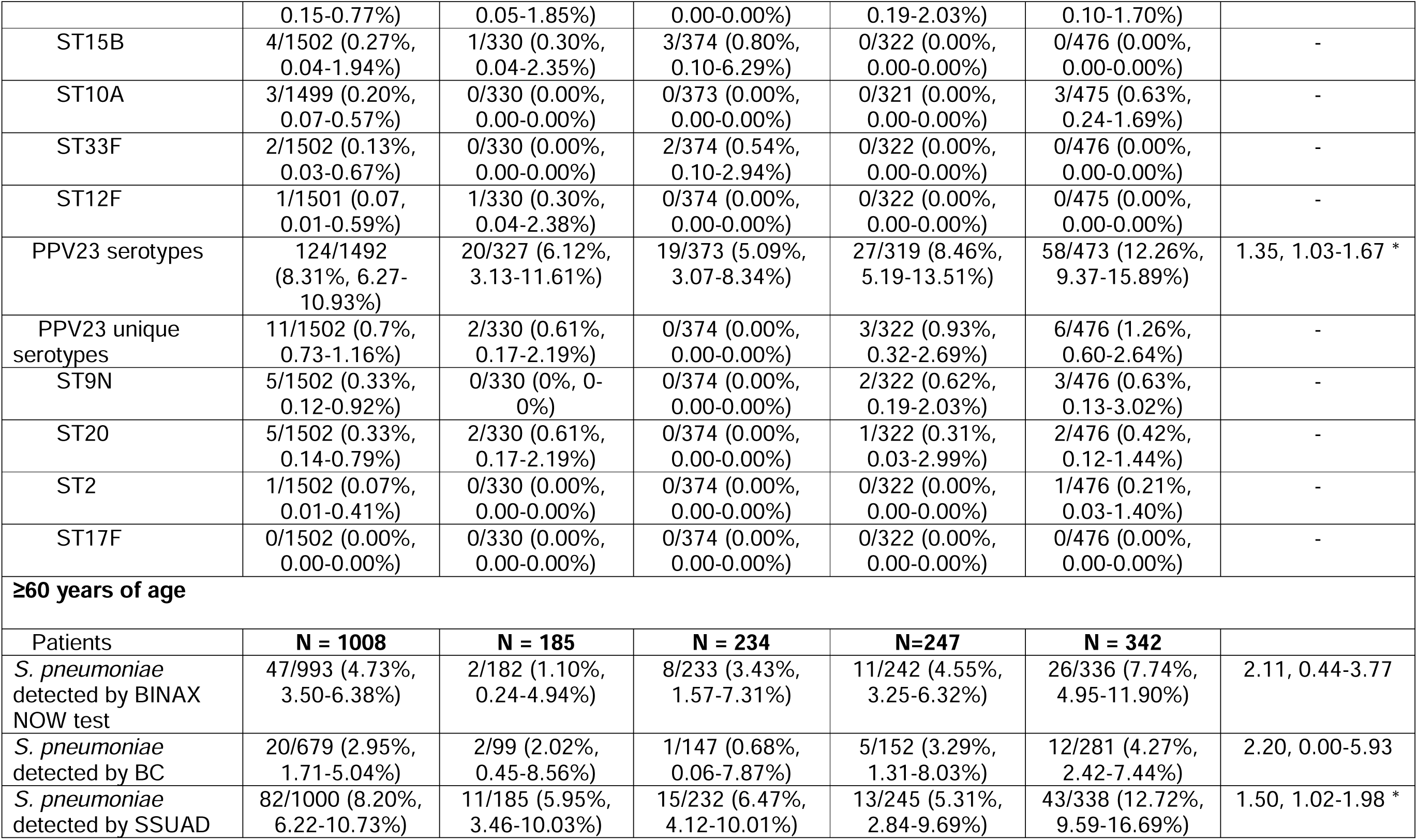
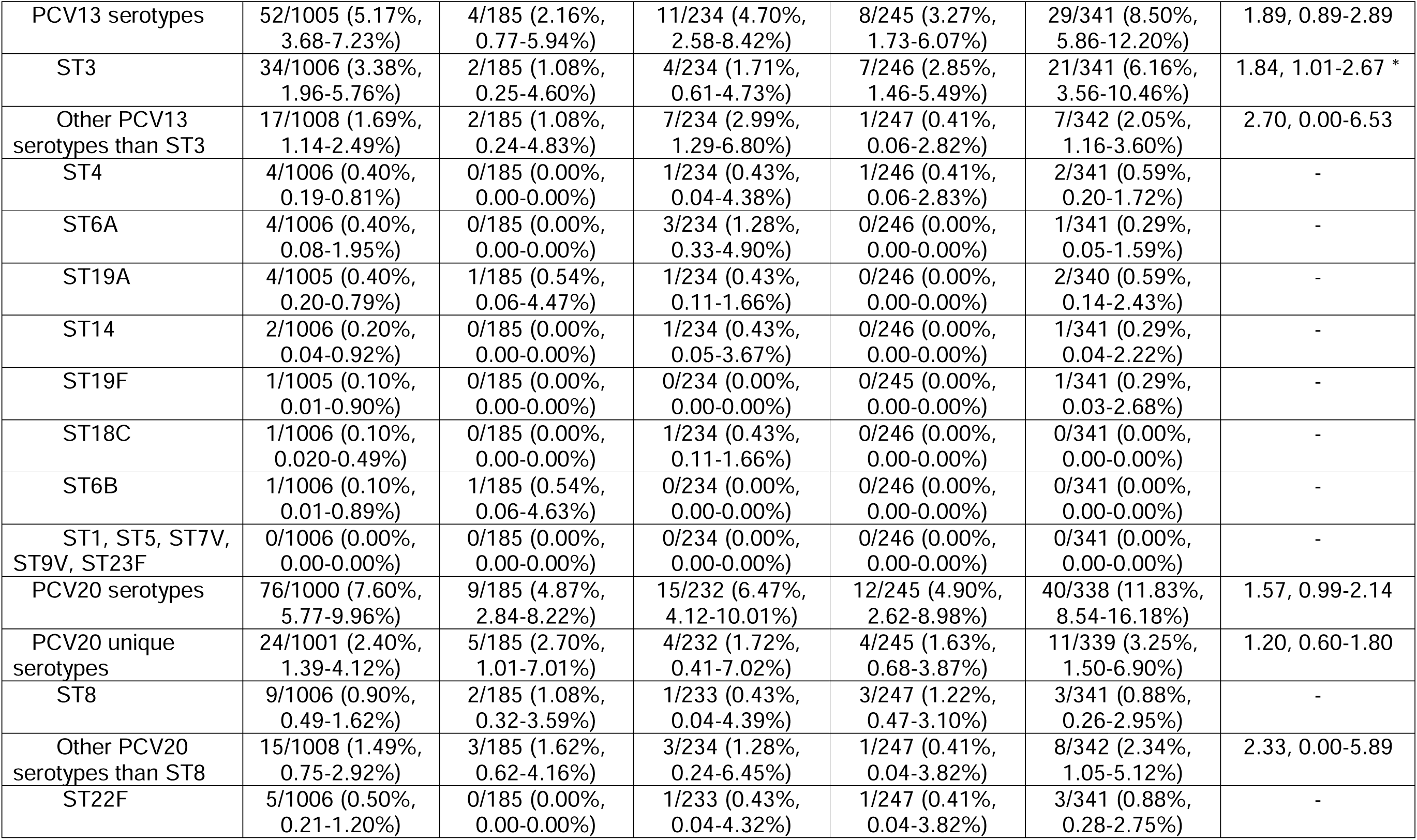
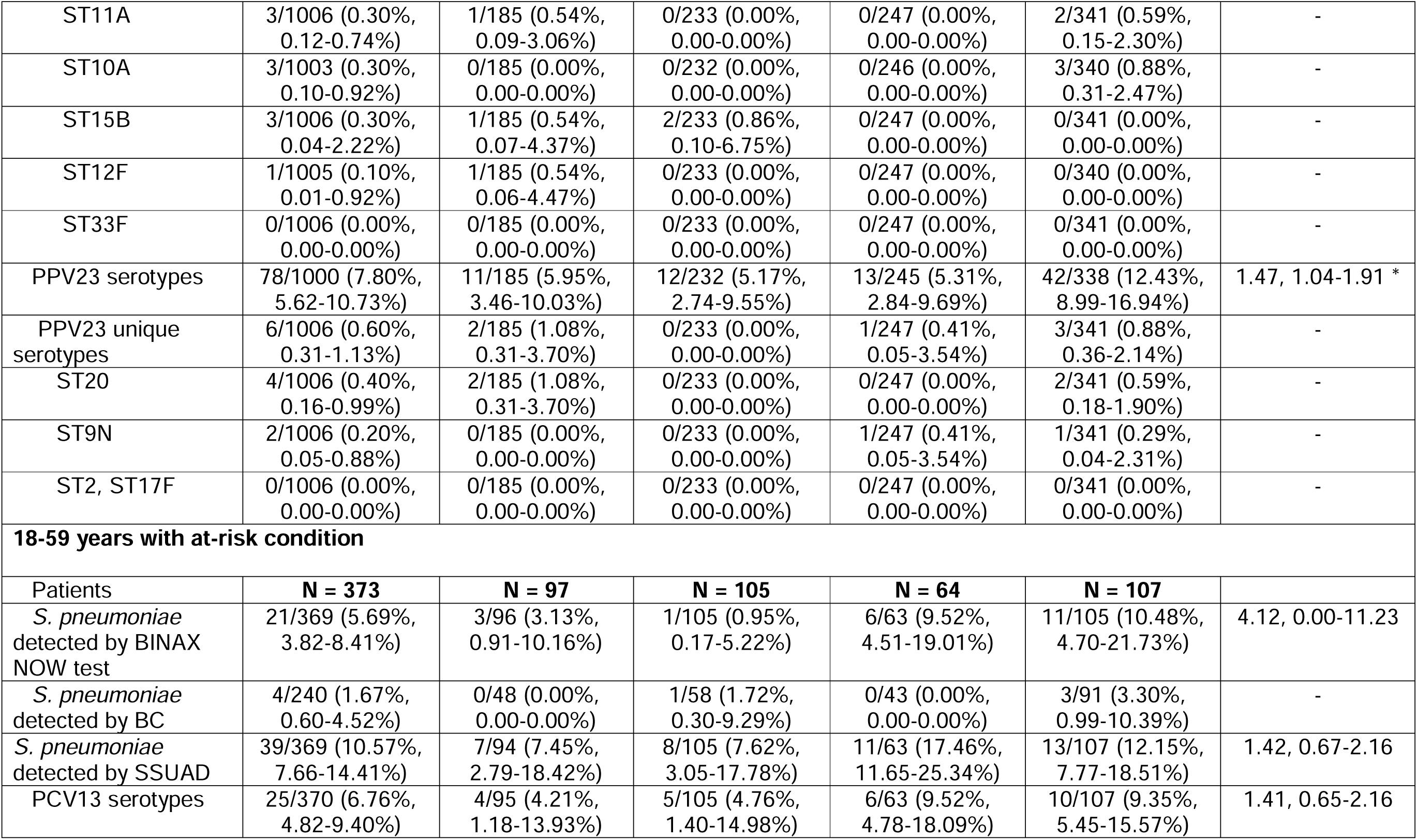
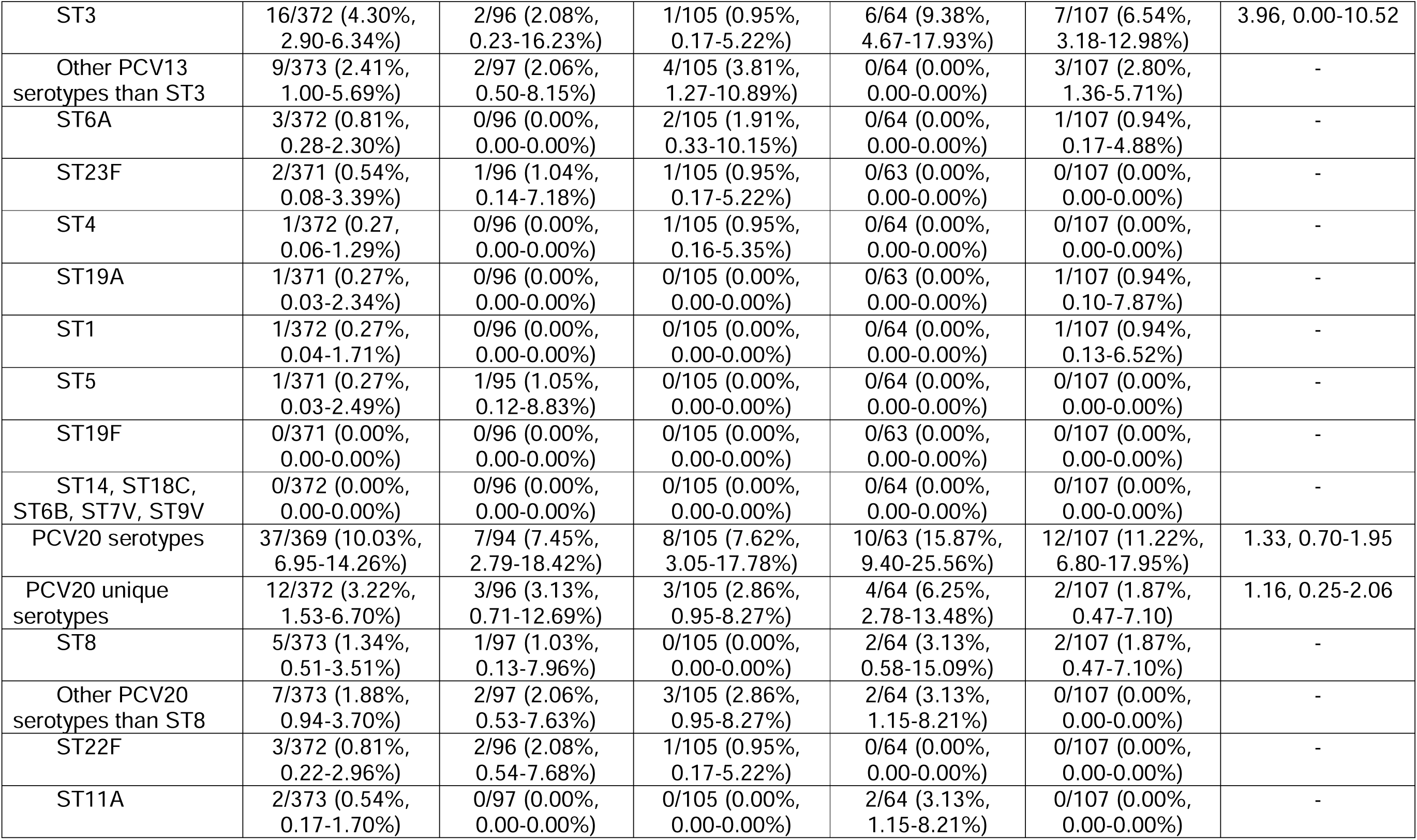
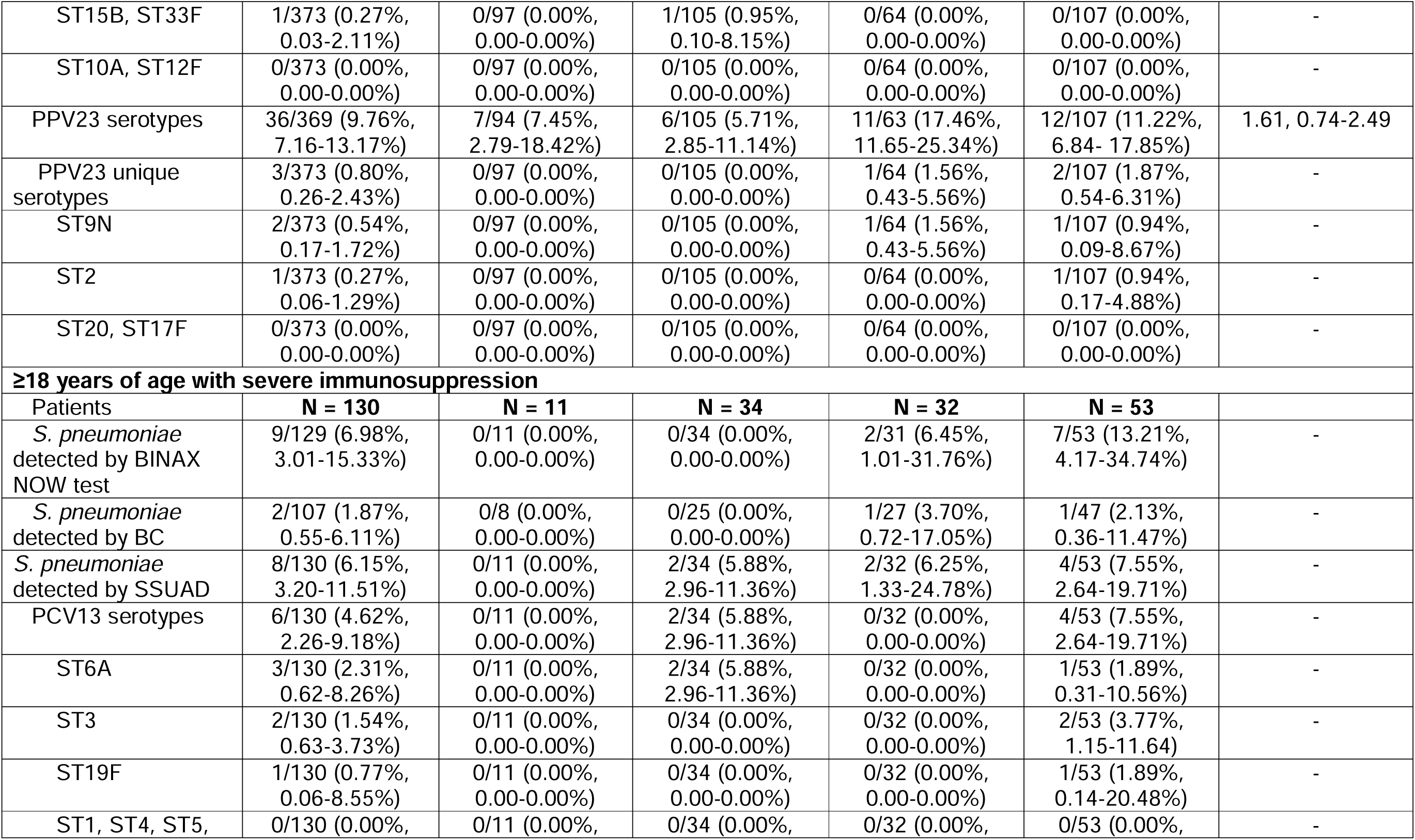
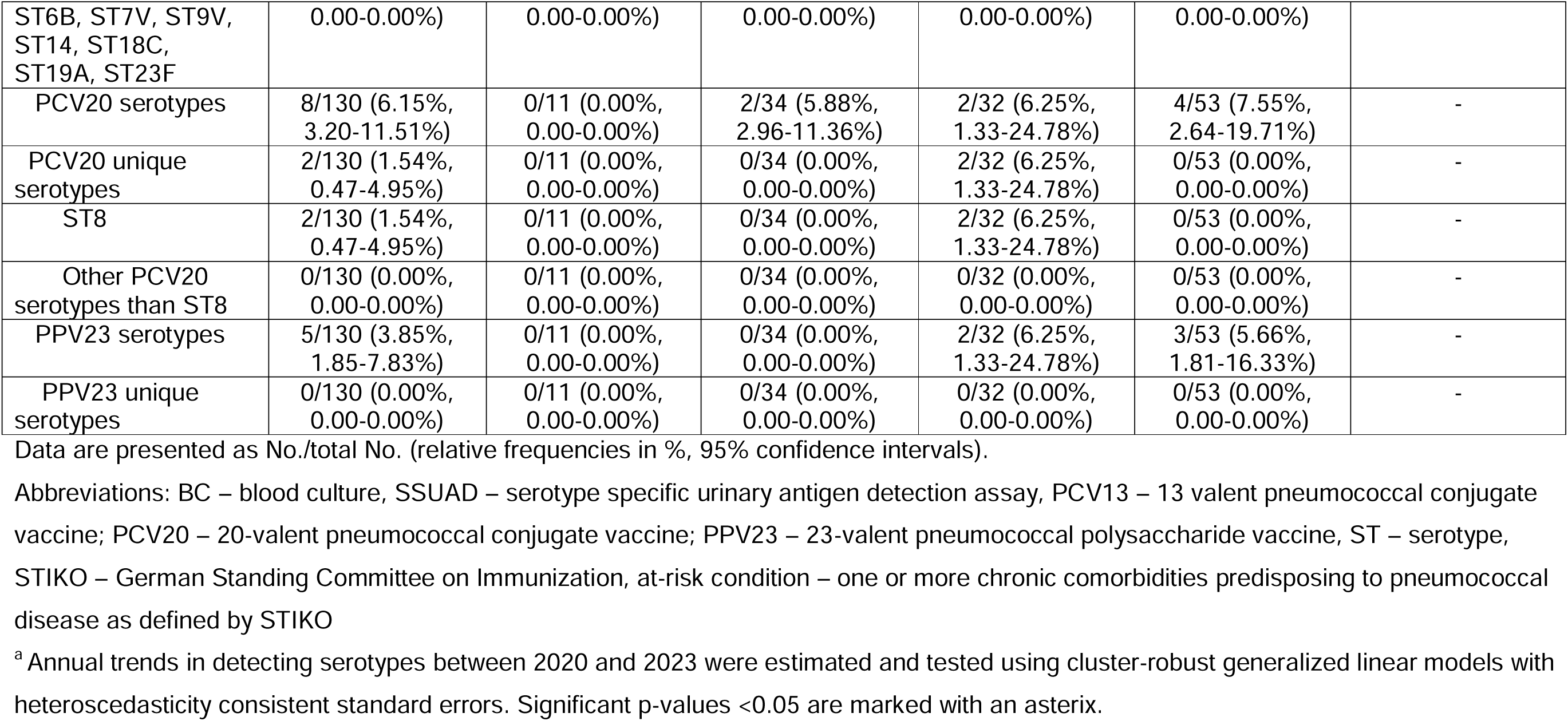
*Streptococcus pneumoniae* detected by conventional methods (BINAX NOW urine antigen test, blood culture) and distribution of pneumococcal vaccine serotypes detected by SSUAD in adult patients with community-acquired pneumonia and in patient subgroups per STIKO recommendation for pneumococcal vaccination (individuals ≥ 60 years, individuals 18-59 years with at-risk condition, i.e. ≥1 comorbidity, individuals ≥18 years with severe immunosuppression)

An important limitation is that SSUAD does not cover 8 additional serotypes (15A, 15C, 16F, 23A, 23B, 24F, 31, 35B) included in the new pneumococcal conjugate vaccine Capvaxive (V116) [11].

These results should be interpreted in light of the national vaccination coverage in Germany (https://www.rki.de/vacmap): During the study period, pneumococcal vaccination rates (≥1 dose) were 11.3–13.2% among 60–64-year-olds and 38.9– 40.2% among 70–74-year-olds. National surveillance does not distinguish between conjugate vaccines and PPV23; however, PPV23 remained the standard adult vaccine until 2023 and likely accounts for the majority of vaccinations.

In conclusion, *S. pneumoniae* has reemerged as a leading CAP-pathogen in adults, particularly among individuals aged ≥60 years in the CAPNETZ patient cohort. This resurgence during the SARS CoV-2 pandemic has been primarily driven by an increase in cases of serotype 3 (1.08% of all-cause CAP in 2020 versus 6.16% of all-cause CAP in 2023). In 2023, PCV20 still showed a high coverage of all-cause CAP in adults (11.83% for age group ≥60 years and 11.22% for age-group 18-59 years with ≥1 comorbidity), reaching prepandemic levels in Germany [7]. Our data show i) an increase of serotype 3 and PPV23 serotypes among all-cause CAP cases between 2020-2023 and ii) that the gap in the coverage between PCV20 and PPV23 remains small.

## Data Availability

All data produced in the present study are available upon reasonable request to the authors

## Abbreviation list

BC: blood culture
CAP: community-acquired pneumonia
CI: confidence interval
OR: odds ratio
PCV13: 13-valent conjugate vaccine
PCV20: 20-valent conjugate vaccine
PPV23: 23-valent pneumococcal polysaccharide vaccine
STIKO: German Standing Committee on Immunization
SSUAD: serotype-specific multiplex urinary antigen detection assay

## Statements

### Data availability

The data sets used and/or analysed during the current study are available upon reasonable request from the corresponding author.

### Ethical statement

Research was conducted in accordance with the Declaration of Helsinki and national and institutional standards. The study protocol was approved by the local ethics committees of all participating centres (number of leading ethics committee Medical Faculty of Otto-von-Guericke-University Magdeburg: 104/01; see acknowledgment or www.capnetz.de for participating centres).

Prior to study enrolment, all patients provided written informed consent. As only anonymised data were analysed, additional consent to publish was not required.

## Acknowledgements

CAPNETZ is a multidisciplinary approach to better understand and treat patients with community-acquired pneumonia. Members of the CAPNETZ study group are: A. Fuchs, G. Paul, M. Ayoub (III. Medical Clinic, University Hospital Augsburg); A. Prasse (Clinic of Pneumology, University Hospital Basel, Switzerland); W. Bauer, E. C. Diehl-Wiesenecker, N. Galtung (Central Emergency Admission / Medical Admission Ward, Charité-Universitätsmedizin Berlin), N. Suttorp, M. Witzenrath, C. Kodde, Y.-M. Stoppe (Clinic of Pneumology, Respiratory Medicine and Intensive Care Medicine with Study Area of Sleep Medicine, Charité-Universitätsmedizin Berlin); C. Boesecke, S. Breitschwerdt, D. Benke (Medical Clinic and Polyclinic I – General Internal Medicine, University Hospital Bonn); S. Schmager (Pneumology Section of II. Medical Clinic, Carl-Thiem Hospital Cottbus); A. Grünewaldt, J. Wheeler (Department of Pulmonology and Intensive Care Medicine, Alice Hospital Darmstadt; B. Schaaf, J. Kremling (Pneumology, Infectiology and Internal Intensive Care Medicine, Medical Clinic Nord, Dortmund); M. Kolditz, B. Schulte-Hubbert, J. Ronczka (Medical Clinic I / Department of Pneumology, University Hospital Dresden); G. Rohde, A. Seeger, J. Kohlhäufl (Medical Clinic I - Pneumology/Allergology, Hospital of Johann Wolfgang Goethe University, Frankfurt), D. Stolz, S. Fähndrich (Clinic of Pneumology, University Hospital Freiburg), M. Panning (Institute of Virology, University Hospital Freiburg); M. Unnewehr, R. Lim (Department of Internal Medicine V: Pulmonology, Infectiology, Sleep Medicine, Allergology, St. Barbara-Clinic Hamm-Heessen); M. Hoeper, I. Pink, N. Drick (Department of Pneumology, Hannover Medical School, Hannover), T. Fühner, T. Steinberg (Clinic of Pneumology, Intensive Care and Sleep Medicine, Siloah Hospital, Hannover), G. Barten-Neiner, W. Kröner, O. Unruh, N. Adaskina, F. Eberhardt, G. Krishnamoorthy (CAPNETZ Office, Hannover), T. Illig, N. Klopp (Hannover Unified Biobank, Hannover Medical School); M. Pletz, B. T. Schleenvoigt, A. Moeser (Institute for Infection Medicine and Hospital Hygiene (IIMK), University Hospital Jena); D. Drömann, P. Parschke, K. Franzen (Medical Clinic III, Pneumology, University Hospital Schleswig-Holstein, Lübeck), J. Rupp, F. Kadgien, B. Gebel, N. Käding, S. Boutin (Clinic and Polyclinic of Internal Medicine II, Infectiology, University Hospital of the Technical University of Munich); J. Schneider, J. Erber, F. Voit, (Clinic and Polyclinic of Internal Medicine II, Infectiology, University Hospital of the Technical University of Munich); D. Heigener, I. Hering (Department of Pneumology, Agaplesion Diakonieklinikum Rotenburg); W. Albrich, F. Rassouli, B. Wirth (Clinic of Infectious Diseases, Infection Prevention and Travel Medicine, HOCH Health Eastern Switzerland, Cantonal Hospital St. Gallen, Switzerland); C. Neurohr (Department of Pneumology and Respiratory Medicine, Robert Bosch Hospital, Lung Center Stuttgart); A. Essig, S. Stenger (Institute of Medical Microbiology and Hygiene, University Hospital Ulm), M. Wallner (2mt Software, Ulm); H. Burgmann, L. Traby, L. Schubert (University Clinic of Internal Medicine I, Medical University of Vienna); and all study nurses.

## Author’s contribution and guarantor statement

All authors have made substantial contribution to the study design, data collection, analysis or interpretation, drafting the article and revising it critically for important intellectual content. All authors approved the final version to be submitted. GB-N, MWP, GR, MW, JR, CB designed the study, CB and MWP drafted the article, CB and NR performed the statistical analysis. CB, NR, CF-S, GB-N, MPGL, GR, JR, MW, JeRa, MWP contributed to the critical revisions, and final approval of the article.

